# Burden of Multiple Sclerosis in Eastern Mediterranean Region (1990-2016): Findings From the 2016 Global Burden of Disease Study

**DOI:** 10.1101/19005918

**Authors:** Mohammad Ali Sahraian, Pouria Heydarpour, Maziar Moradi-Lakeh, Sharareh Eskandarieh, Seyed-Mohammad Fereshtehnejad, Kia Vosughi, Vafa Rahimi-Movaghar, Farshad Farzadfar, Reza Malekzadeh, Theo Vos, Valery Feigin, Mohsen Naghavi, Ali H. Mokdad

## Abstract

**Background:** Multiple sclerosis (MS) is among the leading causes of disability in Young Adults worldwide. Current estimates of MS burden in Eastern Mediterranean Region (EMR) are necessary for planning effective interventions .To estimate Prevalence, incidence, mortality, years lived with disability (YLDs), years of life lost (YLLs) and disability-adjusted life-years (DALYs) in EMR by country age, and sex from 1990 to 2016.

**Methods:** We estimated regional and country-specific prevalence, incidence, mortality, DALYs, YLLs, and YLDs for MS. DALYs were computed as the sum of YLDs and YLLs.

**Results:** Total DALYs in EMR countries was 12,810 in 1990 for males and increased to 36,391 in 2016 and from 18,962 to 53,851 for females. Lowest DALYs in both sexes were observed in Somalia (248) while the highest were in Iran (26,394). YLDs in males increased from 6,511 in 1990 to 19,515 in 2016, and in females from 12,247 to 33,937. The highest age-standardized prevalence, incidence, YLDs and DALYs were in Iran (72.11, 2.49, 18.03, and 32.5, respectively).

**Conclusions:** Our findings provide valuable information to guide the development and implementation of measures to address the rising burden of MS and it consequences in the EMR countries.

## Background

Multiple sclerosis (MS) is an inflammatory demyelinating disease of central nervous system affecting 2.3 million people worldwide (Browne et al., 2014). Disability-adjusted life-years (DALYs) is a widely used health gap measure which shows differences between a population and a normative standard of maximum lifespan at full health. DALYs is the sum of years of life lost (YLLs) due to premature mortality and years lived with disability (YLDs) (DALYs and Collaborators, 2017).

Global prevalence of MS was 2,221 (2,034-2,437) thousands globally in 2016 with a global incidence of 69 (63-76) thousands. MS mainly affects young adults, thus contributing to 584 (411-764) thousand YLDs globally in 2016 with 24.5 (21.9-26.9) percent increase in counts compared to 2006 (Disease et al., 2017). Global death for all ages from MS in 2016 was 18.9 (16.6-21.0) thousands with 17.1 (5.4-22.5) percent increase compared to 2006 (Mortality and Causes of Death, 2017). All-age DALYs globally was 1,151.5 (968.6-1,345.8) in 2016 increased 18.2 (12.8-21.9) percent compared to 2006 (DALYs and Collaborators, 2017).

Eastern Mediterranean Region (EMR) has a population of 583 million people in countries with varied gross domestic product, health system capacities, surveillance systems, and data availability due to limited epidemiological studies (Kobelt et al., 2017). A meta-analysis on the prevalence of MS in Middle East and North Africa showed an overall prevalence of 51.52 per 100,000 in 2015 (Heydarpour et al., 2015).

Despite overall and cause-specific improvements in mortality of MS patients, respiratory causes of death remain highest cause of excess mortality in MS patients and MS patients have an increased mortality risk at all ages in a population-based cohort with extended follow-up (Burkill et al., 2017). A cost of illness study in Europe revealed that costs of MS was correlated with disease severity (Kobelt et al., 2017). We used results of the Global Burden of Disease (GBD) 2016 study to report MS burden in EMR countries to inform policy makers of the current and trend of MS burden.

## Methods

GBD 2016 covers 195 countries, 21 regions, and seven super-regions from 1990 to 2016 for 328 diseases and injuries, 2,982 sequelae, and 84 risk factors by age and sex. Detailed descriptions of GBD 2016 methodology and neurological disorders methodology have been provided elsewhere (DALYs and Collaborators, 2017; Disease et al., 2017; Factors and Collaborators, 2017; Group, G.N.D.C., 2017; Mortality and Causes of Death, 2017).

We evaluated the MS burden in 22 EMR countries: Afghanistan, Bahrain, Djibouti, Egypt, Iran, Iraq, Jordan, Kuwait, Lebanon, Libya, Morocco, Pakistan, Palestine, Qatar, Saudi Arabia, Somalia, Sudan, Syria, Tunisia, United Arab Emirates (UAE) and Yemen.

All-cause mortality was first estimated for each country during the period of 1990–2016. For this purpose, we used all accessible data from vital registration systems, sibling history surveys, sample registration data, and household recall of deaths. We extracted causes of death data from the same sources, as well as available verbal autopsies, and then used GBD’s Cause of Death Ensemble model to estimate the number of deaths from MS by age, sex, country, and year. In this approach, a large variety of possible models are explored to estimate trends in causes of death. Possible models are identified based on a covariate selection algorithm that yields several plausible combinations of covariates; they are then run through different model classes, including mixed effects linear models and spatiotemporal Gaussian process regression models for cause fractions and death rates. All models for each cause of death are then assessed using out-of-sample predictive validity and combined into an ensemble with optimal out-of-sample predictive performance (Mortality and Causes of Death, 2017).

We updated our previous systematic reviews for the GBD study separately for each of the non-fatal sequelae of MS. Non-fatal burden estimates were based on a systematic review of the literature to obtain all available epidemiological data on prevalence, incidence, risk of mortality, and severity). List of all sources (by cause and location) are available at the Institute for Health Metrics and Evaluation’s website. A list of GBD sequelae, health states, health state lay descriptions, and disability weights for MS is provided (Group, G.B.D.N.D.C., 2017).

Bayesian meta-regression analysis through DisMod-MR 2.1 was used for disease modeling. Model-based prevalence estimates, in combination with disability weights, were used to calculate cause-specific YLDs for each age, sex, location, and year. DALYs were calculated through summation of YLLs and YLDs (DALYs and Collaborators, 2017).

GBD 2015 introduced Socio-demographic Index (SDI) to provide a summary measure of a geography’s socio-demographic development. It is based on average income per person, educational attainment, and total fertility rate. In GBD 2015, SDI was computed by rescaling each component to the scale of zero to one, and then taking the geometric mean of these values for each location-year. Zero indicates the lowest observed educational attainment, lowest income per capita, and highest fertility rate from 1980 to 2015, and one indicates the highest observed educational attainment, highest income per capita, and lowest fertility rate during that time. The global SDI in 2015 was 0.638, and the SDI in this region ranged from 0.150 in Somalia to 0.874 in United Arab Emirates (2015., 2016).

We report 95% uncertainty intervals (UI) for each estimate, including rates, numbers of deaths, and DALYs. We estimated UIs by taking 1000 samples from the posterior distribution of each quantity and using the 25th- and 975th-ordered draws of the uncertainty distribution.

## Results

### Prevalence

Highest all age cases of MS in EMR countries in 2016 was estimated to be 58650 (53713-63984), 29780 (26263-33750), and 29566 (26112-33348) in Iran, Pakistan and Egypt respectively. Lowest all age cases of MS in this region in 2016 was estimated as 44 (39-51), 277 (240-318), and 543 (477-620) cases in Djibouti, Somalia and Bahrain. Highest all age cases of female patients was estimated to be 36643 (33397-40293), 20020 (17497-22683), and 19577 (17170-22151) in Iran, Egypt and Pakistan.

Highest all age prevalence rate of MS in EMR countries was estimated as 72.11 (66.04-78.67), 58.82 (52.10-66.87), and 54.08 (48.07-61.37) per 100,000 in Iran, Lebanon, and Tunisia in 2016. Lowest all age prevalence rates in this region was estimated to be 2.66 (2.30-3.06), 4.56 (3.98-5.25), and 13.05 (11.55-14.89) per 100,000 in Somalia, Djibouti and Yemen. Highest all age prevalence female to male ratio was observed in Jordan (2.33), Morocco (2.17), and Yemen (2.16) and lowest ratio was observed in UAE (1.63), Oman (1.68), and Iran (1.69).

Highest age-standardized prevalence rate of MS in EMR countries was estimated to be 70.22 (64.81-76.35), 57.98 (51.44-65.90), 50.77 (45.11-57.50) per 100,000 in Iran, Lebanon and Tunisia in 2016. Lowest age-standardized prevalence rate of MS was estimated as 4.07 (3.53-4.65), 5.94 (5.22-6.78), 19.41 (17.15-21.90) per 100,000 in Somalia, Djibouti, and Pakistan. Highest age-standardized female to male ratio was observed in Jordan (2.33), UAE (2.26), and Saudi Arabia (2.20) and lowest ratio was observed in Iran (1.66), Palestine (1.93), and Afghanistan (1.95). All age prevalence rates and female to male ratios and age-standardized prevalence rates and female to male ratios for EMR countries in 1990 and 2016 are shown in Table 1. Prevalence rates for various age groups for Iran is depicted in online supplementary figure 1.

### Incidence

Highest age-standardized incidence rate in EMR countries for MS was estimated as 2.49 (2.27-2.74), 1.89 (1.67-2.17), 1.65 (1.47-1.87) per 100,000 in Iran, Lebanon, and Tunisia in 2016. Lowest age-standardized incidence rate in this region was estimated as 0.19 (0.16-0.21), 0.25 (0.22-0.29), 0.69 (0.60-0.78) per 100,000 in Somalia, Djibouti, and Yemen. Highest age-standardized incidence rate for MS in females was estimated as 2.93 (2.65-3.24), 2.43 (2.14-2.78), 2.21 (1.94-2.52) per 100,000 in Iran, Lebanon, and Jordan. Lowest age-standardized incidence rate for MS in females was estimated as 0.24 (0.21-0.27), 0.32 (0.28-0.37), 0.88 (0.77-1.00) per 100,000 in Somalia, Djibouti, and Yemen. Highest age-standardized incidence rate for MS in males was estimated as 2.05 (1.83-2.27), 1.30 (1.15-1.48), 1.19 (1.07-1.36) per 100,000 in Iran, Lebanon, and Tunisia. Lowest age-standardized incidence rate in males was estimated as 0.13 (0.12-0.15), 0.18 (0.16-0.21), 0.50 (0.44-0.57) per 100,000 in Somalia, Djibouti, and Yemen.

Age-standardized incidence rate per 100,000 for MS in female and male patients in EMR countries in 1990 and 2016 are shown in Figure 1A, and 1B.

**Figure 1.**
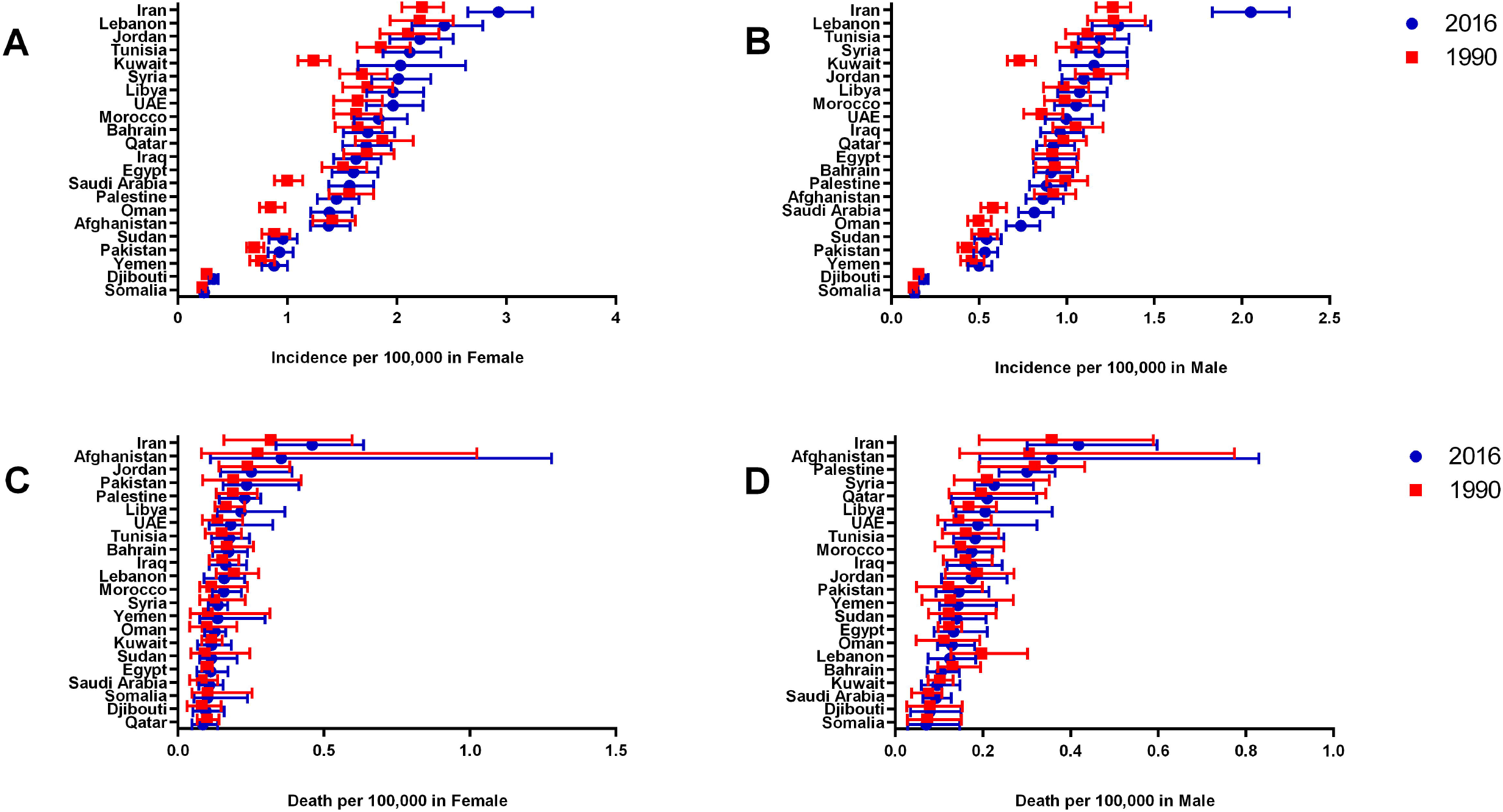
Age-standardized incidence and mortality rate per 100,000 for Multiple Sclerosis in female and male patients in EMR countries in 1990 and 2016.

### Mortality

Highest age-standardized death rate in EMR countries for MS was estimated to be 0.44 (0.35-0.57), 0.35 (0.18-0.75), 0.26 (0.20-0.30) per 100,000 in Iran, Afghanistan and Palestine in 2016. Lowest age-standardized death rate in this region was estimated as 0.086 (0.05-0.15), 0.087 (0.05-0.14), 0.10 (0.08-0.13) per 100,000 in Somalia, Djibouti, and Saudi Arabia. Highest age-standardized death rate in EMR countries for MS in females was estimated as 0.46 (0.34-0.64), 0.35 (0.11-1.28), 0.25 (0.15-0.39) per 100,000 in Iran, Afghanistan, and Jordan in 2016. Lowest age-standardized death rate in this region for MS in females was estimated as 0.09 (0.05-0.14), 0.095 (0.05-0.16), 0.10 (0.05-0.24) per 100,000 in Qatar, Djibouti, and Somalia. Highest age-standardized death rate for MS in males was estimated as 0.42 (0.30-0.60), 0.36 (0.19-0.83), 0.30 (0.24-0.36) per 100,000 in Iran, Afghanistan, and Palestine. Lowest age-standardized death rate for MS in males was estimated as 0.07 (0.03-0.15), 0.08 (0.03-0.15), 0.09 (0.06-0.13) per 100,000 in Somalia, Djibouti, and Saudi Arabia.

Age-standardized death rate per 100,000 for MS in female and male patients in EMR countries in 1990 and 2016 are shown in Figure 1C, and 1D.

### YLDs

Highest age-standardized YLDs rate in EMR countries for MS was estimated as 18.03 (12.73-23.49), 14.96 (10.41-19.74), 13.19 (9.12-17.70) per 100,000 in Iran, Lebanon, and Tunisia in 2016. Lowest age-standardized YLDs rate for MS in this region was estimated as 1.16 (0.79-1.57), 1.70 (1.14-2.28), 5.30 (3.54-7.09) per 100,000 in Somalia, Djibouti, and Yemen. Highest age-standardized YLDs rate in female MS patients was estimated as 22.29 (15.26-29.46), 20.28 (14.01-27.44), 18.18 (12.23-24.35) per 100,000 in Iran, Lebanon, and Jordan. Lowest age-standardized YLDs rate in female patients was estimated as 1.54 (1.05-2.09), 2.26 (1.53-3.05), 6.93 (4.58-9.48) per 100,000 in Somalia, Djibouti, and Yemen. Highest age-standardized YLDs rate in male patients was estimated as 13.81 (9.64-18.58), 9.84 (6.64-13.40), 9.09 (5.97-12.78) per 100,000 in Iran, Lebanon, and Kuwait. Lowest age-standardized YLDs rate in male patients was estimated as 0.77 (0.52-1.05), 1.13 (0.76-1.53), 3.63 (2.47-4.89) per 100,000 in Somalia, Djibouti, and Yemen.

Age-standardized YLDs rate per 100,000 for MS in female and male patients in EMR countries in 1990 and 2016 are shown in Figure 2A, and 2B.

**Figure 2.**
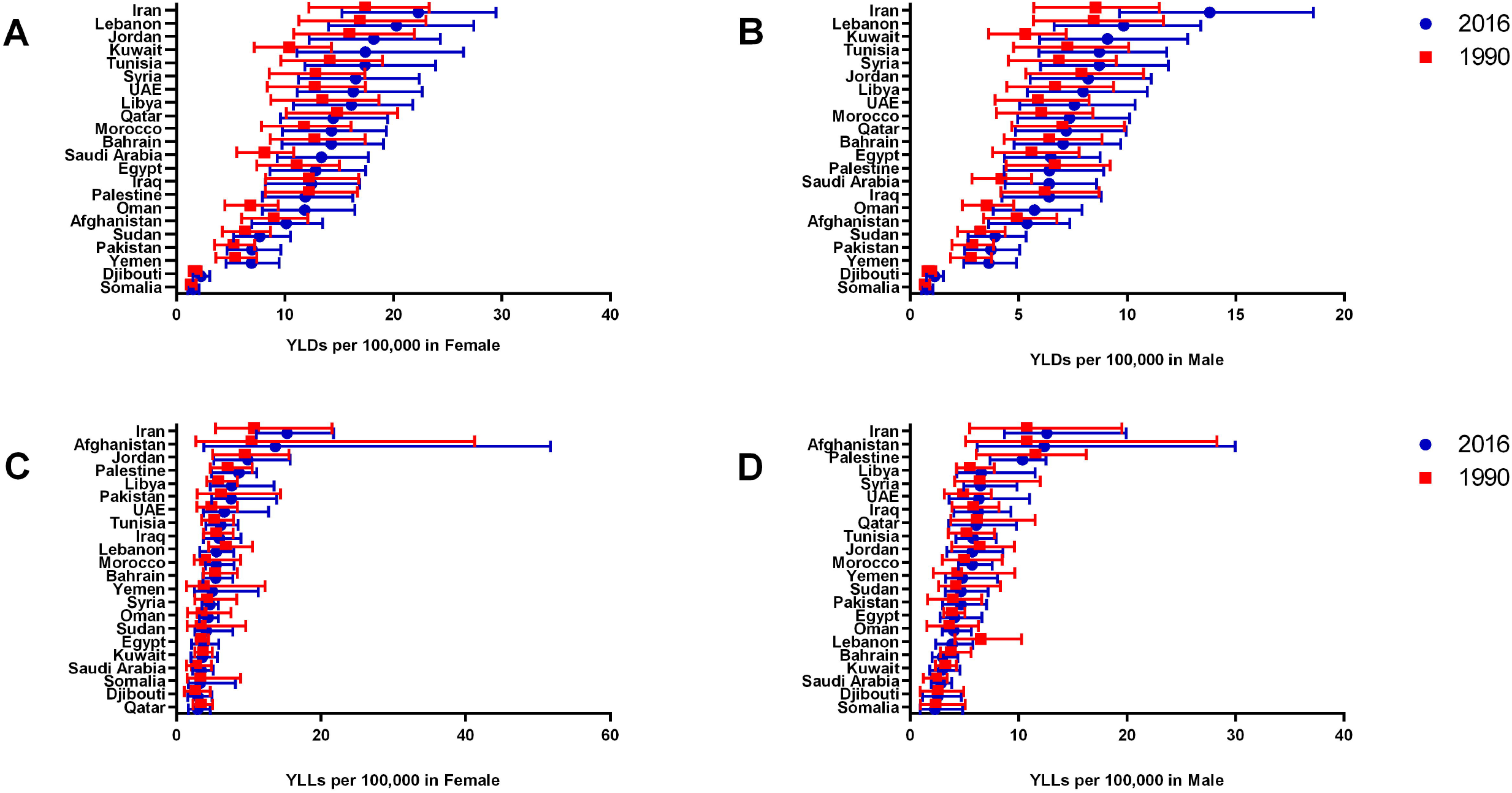
Age-standardized YLDs and YLLs rate per 100,000 for Multiple Sclerosis in female and male patients in EMR countries in 1990 and 2016.

### YLLs

Highest age-standardized YLLs rate for MS in EMR countries was estimated as 13.94 (10.89-19.38), 12.96 (6.15-29.51), 9.48 (6.44-11.34) per 100,000 in Iran, Afghanistan, and Palestine in 2016. Lowest age-standardized YLLs rate for MS in this region was estimated as 2.75 (1.52-4.47), 2.80 (1.70-4.91), 3.08 (2.58-4.05) per 100,000 in Djibouti, Somalia, and Saudi Arabia. Highest age-standardized YLLs rate for female MS patients was estimated as 15.31 (11.05-21.77), 13.70 (3.83-51.75), 9.88 (5.18-15.76) per 100,000 in Iran, Afghanistan, and Jordan. Lowest age-standardized YLLs rate for female MS patients was estimated as 2.94 (1.68-4.71), 2.96 (1.59-4.93), 3.28 (1.70-8.16) per 100,000 in Qatar, Djibouti, and Somalia. Highest age-standardized YLLs rate for male MS patients was estimated as 12.62 (8.70-19.91), 12.37 (6.19-29.97), 10.36 (7.36-12.53) per 100,000 in Iran, Afghanistan, and Palestine. Lowest age-standardized YLLs rate for MS was estimated as 2.28 (0.95-4.85), 2.53 (1.15-4.75), 2.85 (1.94-3.84) per 100,000 in Somalia, Djibouti, and Saudi Arabia.

Age-standardized YLLs rate per 100,000 for MS in female and male patients in EMR countries in 1990 and 2016 are shown in Figure 2C, and 2D.

### DALYs

All age DALYs rate and age-standardized DALYs rate for EMR countries for male and female patients in 1990 and 2016 are shown in Table 2. Highest all age total number of DALYs for MS in EMR countries in 2016 was estimated to be 26395 (20990-32645), 17133 (13630-21638), 10803 (8124-13681) in Iran, Pakistan, and Egypt respectively. Lowest all age total number of DALYs for MS in this region was estimated as 32 (23-45), 197 (149-253), 249 (181-392) in Djibouti, Bahrain, and Somalia. Lowest all age DALYs of MS in female patients was observed in Djibouti (19, 13-26), Bahrain (103, 74-135), and Qatar (112, 77-151).

Highest all age DALYs rate for MS in EMR countries was estimated as 32.5 (25.8-40.1), 20.5 (15.9-25.4), 20 (15.1-25.3) per 100,000 in Iran, Tunisia, and Lebanon in 2016. Lowest all age DALYs rate for MS in this region was estimated to be 2.4 (1.7-3.8), 3.3 (2.4-4.6), 6.6 (4.9-8.9) per 100,000 in Somalia, Djibouti, and Yemen. Highest all age DALYs rate in female patients was observed in Iran (38.8, 29.6-48.7), Lebanon (25.8, 18.8-33.4), and Tunisia (25.6, 19.1-33.0) per 100,000.

Highest age-standardized DALYs rate for MS in EMR countries was estimated as 32.0 (25.5-38.9), 20.8 (15.6-26.6), 20.7 (13.0-37.3) per 100,000 in Iran, Jordan, and Afghanistan. Lowest age-standardized DALYs rate for MS in this region was estimated to be 4.0 (2.8-6.1), 4.4 (3.1-6.3), 10.2 (7.7-13.7) per 100,000 in Somalia, Djibouti, and Yemen. Highest age-standardized DALYs rate in female patients was observed in Iran (37.6, 29.0-46.7), Jordan (28.1, 20.5-36.8), and Lebanon (25.8, 18.8-33.3) per 100,000. Lowest age-standardized DALYs rate in female patients was observed in Somalia (4.8, 3.1-9.9), Djibouti (5.2, 3.6-7.4), and Sudan (11.8, 8.5-16.3) per 100,000. Highest age-standardized DALYs rate in male patients was observed in Iran (26.43, 20.60-34.59), Afghanistan (17.75, 10.74-35.62), Palestine (16.78, 12.99-20.06) per 100,000. Lowest age-standardized DALYs rate in male patients was observed in Somalia (3.05, 1.65-5.52), Djibouti (3.66, 2.22-5.86), and Pakistan (8.42, 6.19-11.02).

Total DALYs in EMR countries was 12,810 in 1990 for males and increased to 36,391 in 2016 and from 18,962 to 53,851 for females. Lowest DALYs in both sexes were observed in Somalia (248) while the highest were in Iran (26,394). Figure 3A shows MS prevalence and Figure 3B shows YLDs in 5 year age groups in EMR countries, highest prevalence and YLDs in 2016 in females was in 40-44 age group and in males in 45-49 age group. In 1990 highest prevalence and YLDs in females was reported in 35-39 age group and in males in 40-44 age group. Figure 3C shows YLLs in 5 year age groups in EMR countries, highest YLL in females in 2016 was reported in 45-49 age group and in 50-54 age group in males. In 1990, highest YLLs in females were also observed in 45-49 age group while in male was in 40-44 age group. Highest DALYs in female in 2016 was reported in 45-49 age group and in 40-44 age group in males. In 1990, also the highest DALYS in females were reported in 45-49 age group, and also in 40-44 age group in males.

**Figure 3.**
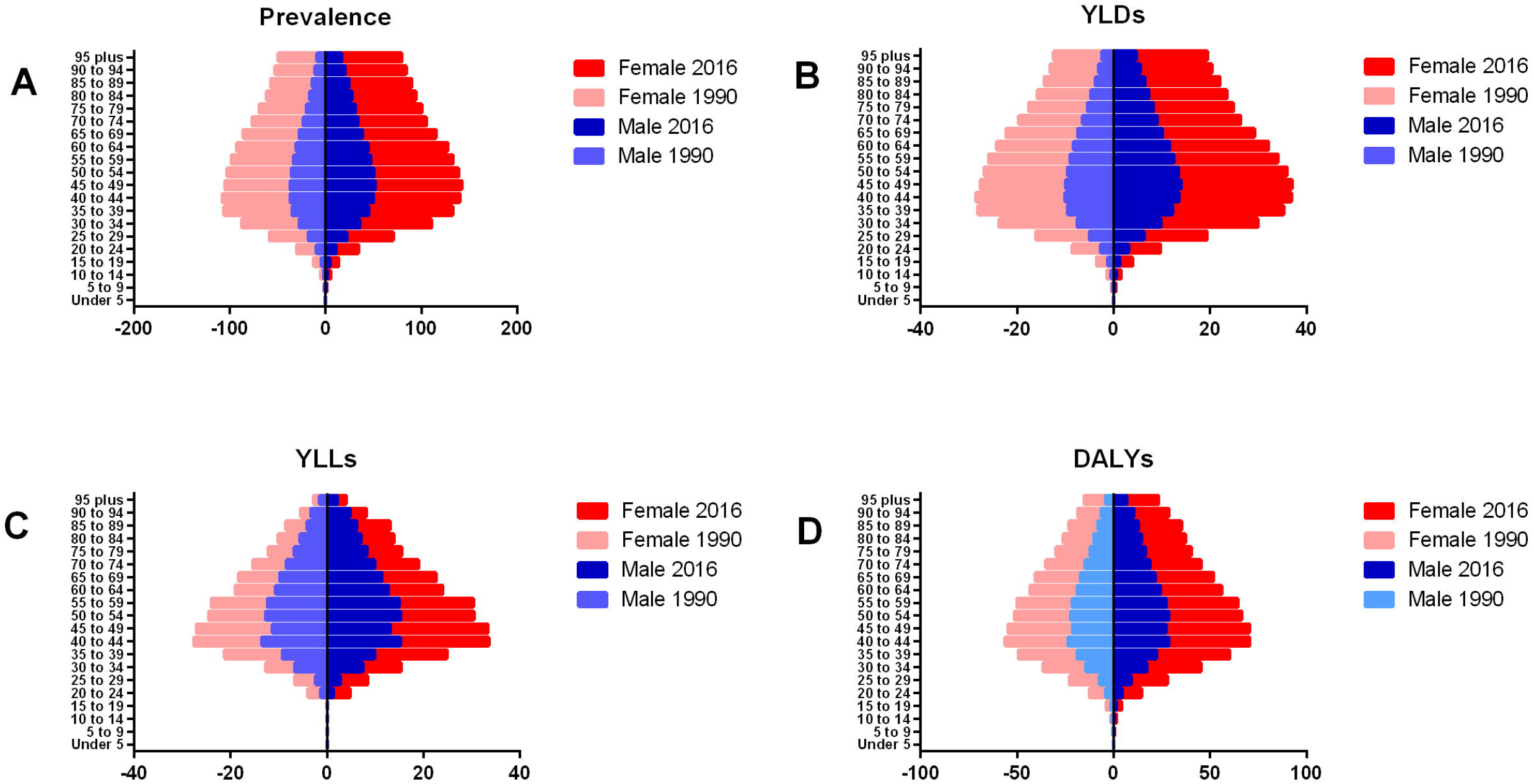
Prevalence, YLDs, YLLs, and DALYs for Multiple Sclerosis in 5 year age groups in female and male patients in EMR countries in 1990 and 2016.

Online supplementary file 2 shows highest DALYs per 100,000 in European region followed by Americas and EMR during 1990 to 2016. As depicted in Figure 4 the highest DALYs annual percent change in 1990-2016 in this region was observed in Saudi Arabia, Iran and Oman. The ratio of observed to expected DALYs ratio (O-E ratio) in 1990 varied from 0.55 in Djibouti to 3.21 in Iran, in 2016 O-E ratio varied from 0.56 in Afghanistan to 3.35 in Yemen. Figure 5A shows the O-E ratio based on SDI in 1990 and Figure 5B is in 2016, there was no correlation between SDI and O-E ratio and the linear regression was not significant.

**Figure 4.**
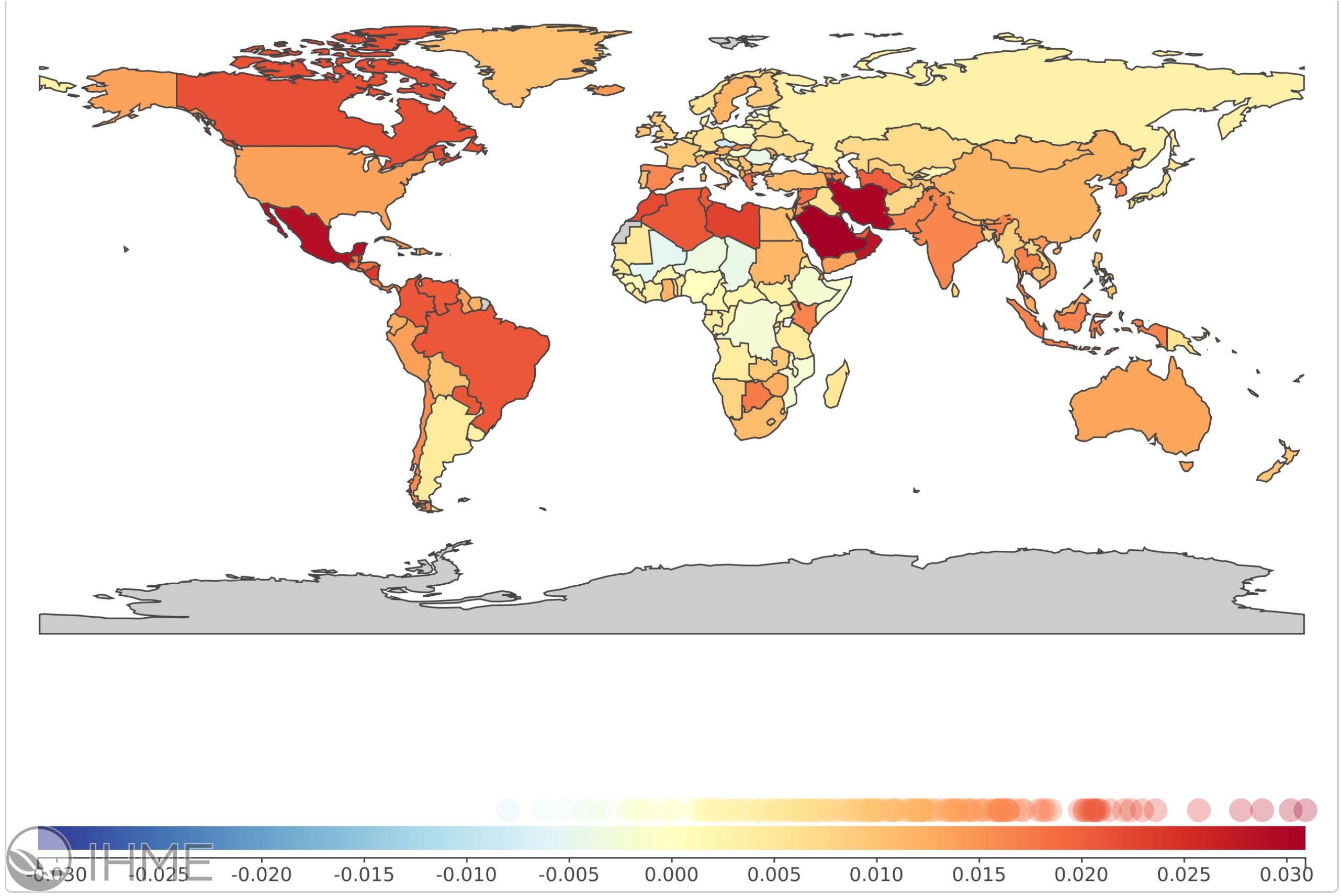
Annual percent change in DALYs in 1990-2016.

**Figure 5.**
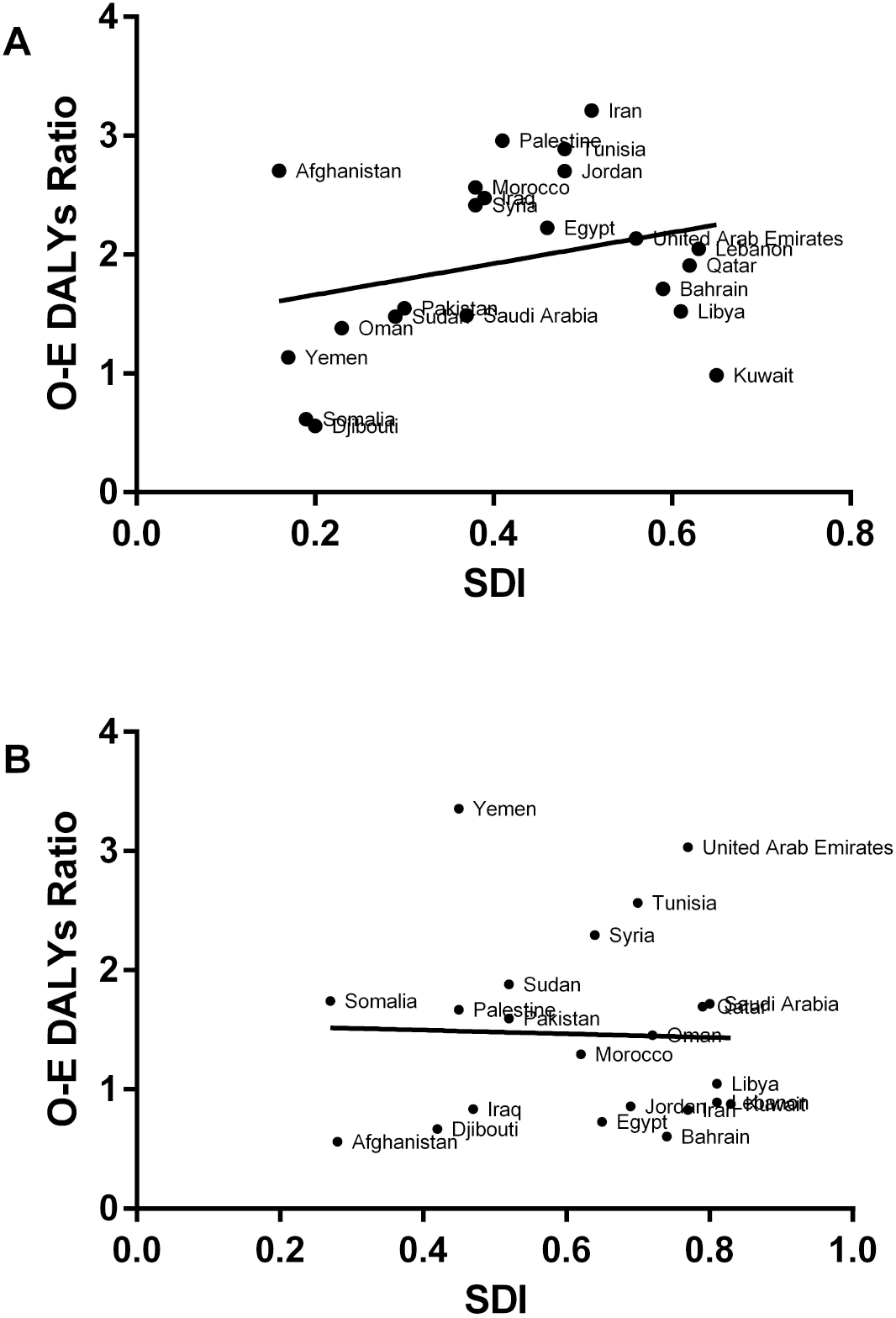
Observed to expected DALYs ratio in 1990 and 2016 based on Socio-demographic Index (SDI).

## Discussion

Our study delivers a comprehensive estimation of prevalence, incidence and DALY of MS among 22 countries in EMR region with important implications for improving health-care planning for people with MS. Our findings are of great importance as they indicated a rapid rise in the burden of MS in the region. With all the health challenges faced in the region, these results should be used to develop and implement programs to reduce the MS burden and its consequences on resources and health systems in the future.

Our results showed that the burden varied by country and was higher among females. Generally, higher female to male ratios of MS in region must be related to the worldwide increase in incidence of MS among women, a similar tendency was observed in Canada and Persian Gulf countries over the last few years (Mohammed, 2016; Orton et al., 2006).

We report the highest and lowest prevalence rate in Iran and Somalia, respectively in 2016. Other studies show in capital city of Iran, MS prevalence had increased to 101.39/100,000 populations in 2014 from 51.9/100,000 in 2008 (Eskandarieh et al., 2017; Sahraian et al., 2010). According to Kurtzke’s classification order (Kurtzke, 1966), most of countries in EMR, (15 from 22) considered countries with a high frequency of MS cases (≥30/100,000), whereas only Somalia is a country with a low frequency of MS cases (<5/100,000) (Kurtzke, 1966).

The prevalence and incidence of MS vary in different parts of region; that may be because lack of registry system and limited nationwide epidemiological data which affect the reporting of MS in some Asian countries (Eskandarieh et al., 2016).

Providing nationwide social health insurance and health-care reforms for special target groups can enable patients to access essential health services and decrease disabilities and mortality rate of chronic diseases such as MS. Health-related Sustainable Development Goals (SDG) index is one of predictors of burden of diseases (Sun et al., 2017).

Rising burden of MS in this region also results in higher burden on healthcare personnel and informal caregivers to MS patients thus more resources will be needed in healthcare systems to provide for direct and indirect costs of MS. Governments in this region need to plan for this rise and plan for future rehabilitation needs of these patients. Registries could help better locate individual patients and address their social and healthcare needs with trained professionals accordingly. Training Neurologists in this region will eventually result in earlier detection of disease so preventive measures could be taken to reduce comorbidities and decrease the long-term disability of patients(Al Tahan et al., 2014; Alroughani et al., 2014; Yamout et al., 2015).

High income countries in North America and Western Europe have periodically developed diagnostic and pharmacological treatment guidelines to inform practicing neurologists of newer medications that might help better control MS. Emerging guidelines from EMR countries in recent years have been successful in informing healthcare providers in this region.

During 1990 to 2016, observed over expected for DALYs in the region has decreased for most of the countries except for Yemen, Somalia and UAE.

Limitations of this study might be due to paucity of epidemiological data on MS in some of the countries, while various data points from various cities from some of these countries was available during 1990 to 2016, other countries only had few data point to allow the estimation of outcomes in this study.

## Conclusion

The increasing trend of MS burden and its different pattern from the global trend needs more investigation. It is important that policy makers and health-care providers be aware of these increasing trends for providing adequate services for the growing numbers of patients with MS and also predict for increased burden of MS in future in their countries.

## Data Availability

All data provided by Institute of Health Metrics and Evaluation is available below.
https://figshare.com/articles/Burden_of_MS_in_EMR/7692389

## Online Supplementary Figure Legends

**Figure 1**. Prevalence rate per 100,000 in five year age groups in Iran in 1990, 2000, 2010, and 2016.

**Figure 2**. DALYS per 100,000 in WHO regions.

